# *Plasmodium falciparum multidrug resistance 1* gene polymorphisms associated with outcomes after antimalarial treatment

**DOI:** 10.1101/2024.07.01.24309724

**Authors:** Veronika R. Laird, Mateusz M. Plucinski, Meera Venkatesan, Kelsey A. Rondini, Milijaona Randrianarivelojosia, Mauricette N. Andriamananjara, Hawela Moonga, Deus S. Ishengoma, Arlindo Chidimatembue, Pedro Rafael Dimbu, Adicatou-Laï Adeothy, Abdoul Habib Beavogui, Simon Kariuki, Sam L. Nsobya, Aline Uwimana, Gauthier Mesia Kahunu, Ashenafi Assefa, Ousmane A. Koita, Naomi W. Lucchi, Samaly S. Svigel Souza, Zhiyong Zhou, Leah F. Moriarty, Eric S. Halsey

## Abstract

This study suggests that: 1) patients given AL infected with parasites carrying N86 were statistically more likely to experience a recurrent infection; 2) patients given ASAQ infected with parasites carrying 86Y were statistically more likely to experience a recurrent infection.

**Background:** *Plasmodium falciparum multidrug resistance transporter 1* (*Pfmdr1*) gene mutations are associated with altered response to artemisinin-based combination therapies (ACTs), particularly those containing the partner drugs lumefantrine and amodiaquine (i.e., artemether-lumefantrine [AL] and artesunate-amodiaquine [ASAQ]). Past studies of *Pfmdr1* single nucleotide polymorphisms (SNPs) at codons 86, 184, and 1246 have shown different responses to AL and ASAQ.

**Methods:** To determine whether infection with parasites carrying specific *Pfmdr1* SNPs leads to increased risk of recurrent parasitemia (recrudescent or new infection), data from 4,129 samples from 16 therapeutic efficacy studies from 13 African countries between 2013–2019 were analyzed.

**Results:** Patients treated with AL and infected with parasites carrying *Pfmdr1* N86 were at greater risk of treatment failure than those whose parasites carried 86Y. After treatment with ASAQ, individuals infected with parasites that carried *Pfmdr1* 86Y were more likely to experience a recurrent infection.

**Conclusions:** Our results support prior studies that suggested: 1) patients given AL and infected with parasites carrying N86 were more likely to experience a recurrent infection; 2) patients given ASAQ and infected with parasites carrying 86Y were more likely to experience recurrent infection. These findings suggest that ACT and *Pfmdr1* genotype may influence outcome after *P. falciparum* infection.

## Introduction

The emergence of *Plasmodium falciparum* resistance to antimalarials poses a challenge to global malaria control efforts. World Health Organization (WHO) Guidelines for Malaria recommend the use of artemisinin-based combination therapy (ACT) to treat uncomplicated *P. falciparum* infection (1). Three ACTs used in Africa are artemether-lumefantrine (AL), artesunate-amodiaquine (ASAQ), and dihydroartemisinin-piperaquine (DP) (2). Although polymerase chain reaction (PCR)-adjusted efficacy for each ACT remains high throughout most regions (3), suboptimal ACT efficacy has been reported in Asia (4) and Africa (5–7).

Although ACT resistance has not been demonstrated with any particular *Plasmodium falciparum multidrug resistance transporter 1* single nucleotide polymorphism (SNP) or haplotype, certain SNPs may affect response after treatment with ACTs containing lumefantrine or amodiaquine (8,9). Five unique *Pfmdr1* SNPs, with amino acid changes in codons 86, 184, 1034, 1042, and 1246, have been found in various regions of the world (10). N86, 184F, and D1246 SNPs have been found in post-treatment samples of patients who experienced a recurrent infection after AL treatment (9,11,12). Conversely, studies of SNPs in *Pfmdr1* have suggested that the 86Y and 1246Y mutations may be associated with decreased response to amodiaquine and ASAQ (9,13). Several *in vivo* studies in sub-Saharan Africa have found evidence of selection for particular *Pfmdr1* SNPs at codons 86, 184, and 1246 after treatment with AL (9,14,15). *Pfmdr1* SNPs present post-treatment in new infections are also important to understand because they may impact the post-treatment prophylactic effect of the partner drug (16). Increased copy number of *Pfmdr1* due to gene duplications has also been associated with reduced susceptibility to lumefantrine (10,17).

Sample sizes in individual therapeutic efficacy studies (TESs) are typically too small to statistically power an assessment of the association between parasite genotypes and treatment outcomes (i.e., adequate clinical and parasitological response, recrudescent infection, and new infection). As part of the US President’s Malaria Initiative (PMI), the Centers for Disease Control and Prevention (CDC) and the US Agency for International Development provide support to countries in Africa to perform TESs, including molecular characterization of antimalarial-resistance markers, through the Partnership for Antimalarial Resistance Monitoring in Africa (PARMA) network (18). Established in 2015, this endeavor involves laboratory scientists from endemic countries coming to CDC (Atlanta, Georgia, USA) or, starting in 2022, to a partner lab in Africa to receive advanced laboratory training and perform molecular testing for antimalarial-resistance mutations (18). To examine the association between parasite genotypes and treatment outcomes, *in vivo* antimalarial efficacy and *P. falciparum* molecular marker data from samples analyzed and collected from 16 TESs conducted in 13 countries in Africa were used. Understanding this association is important for detecting changes in long-term trends of *Pfmdr1* SNP prevalence in parasite populations as indicators of changing parasite susceptibility to ACT partner drugs, lumefantrine or amodiaquine. This study had two main objectives to untangle the relationship between parasite *Pfmdr1-*genotypes and treatment outcomes: 1) To assess whether patients infected with parasites carrying specific *Pfmdr1* SNPs have increased risk of recurrent infection; 2) To investigate preferential selection of specific *Pfmdr1* SNPs by ACT type.

## Methods

### Selection and inclusion of data

Data for this pooled analysis were compiled from 16 individual TESs from 40 sites across 13 sub-Saharan African countries between 2013–2019 (Figure 1, Supplementary Table 1, and Supplementary Table 2). The studies were all funded by PMI.

**Figure 1.**
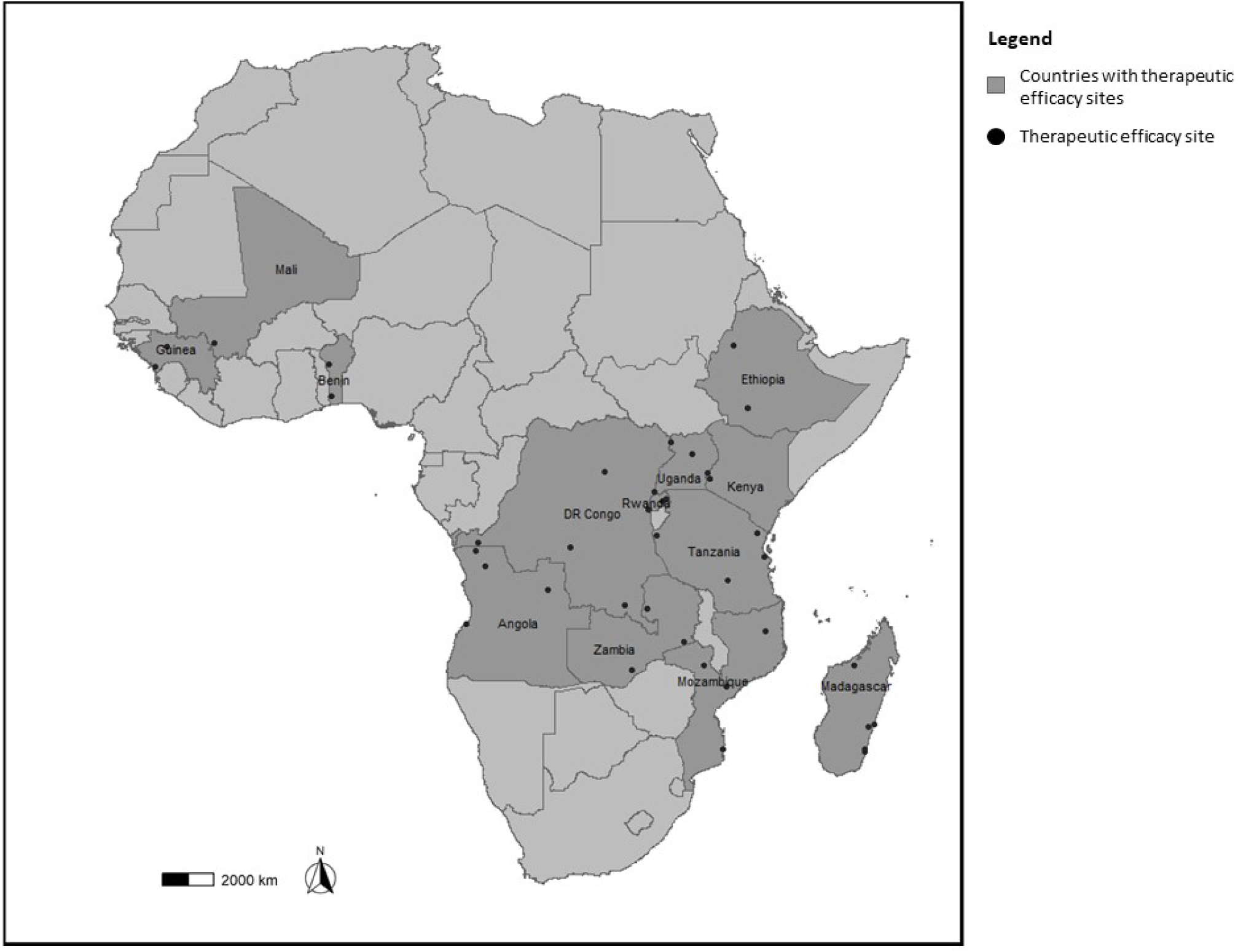
Countries and sites contributing data to this *Pfmdr1* meta-analysis.

Patients with symptomatic, parasitologically-confirmed uncomplicated *P. falciparum* symptomatic infection who presented to health facilities and met standard WHO inclusion criteria were enrolled (Supplementary Table 2) (24). Patients were administered either AL, ASAQ, or DP. After follow-up was complete, parasite genetic sequencing of *Pfmdr1* was conducted at the CDC Malaria Branch Laboratory in Atlanta, USA. This was conducted as part of a training program through PARMA (18). Eleven TES results have been published alongside molecular results, three TES efficacy results were published separately from the molecular surveillance results, and two results are contained in unpublished country reports (Benin and Zambia) (Supplementary Table 2).

### Genotyping of Pfmdr1 SNPs and result calling

Patients were considered to have a recurrent infection if their sample was found by microscopy to contain asexual forms of *P. falciparum* starting on day 4 after ACT treatment until the end of the follow-up period (28, 28, and 42 days for AL, ASAQ, and DP, respectively). For study participants with recurrent infection, the recurrent infection sample and its corresponding pre-treatment sample were genotyped to determine whether the recurrent infection was a recrudescent or new infection. This was done using either seven neutral microsatellite markers (*TA1, Poly-*α*, PfPK2, TA109, TA2490, C2M34*, and *C3M69*) over six chromosomes (20,21) or using the fragment length polymorphism of genes *merozoite surface protein 1*, *merozoite surface protein 2*, and *glutamate-rich protein* (*msp1*, *msp2,* and *glurp*) (22).

*Pfmdr1* codons 86, 184, 1034, 1042, and 1246 were sequenced in parasites collected from enrolled patients who experienced a recurrent infection from each TES and the SNP at each codon was recorded. Each TES sequenced *Pfmdr1* codons 86, 184, 1034, 1042, and 1246 for typically 10% of enrolled patients who experienced an adequate clinical and parasitological response except for the Angola (2013, 2017, 2019), Democratic Republic of the Congo, and Uganda TESs. Sanger sequencing was used to determine the genotype of the five *Pfmdr1* SNPs in pre-treatment and recurrent infections samples for each study except the 2019 Angola TES, which utilized next generation sequencing. For each SNP, samples were classified as having either a single or mixed infection (mixed genotypes) depending on whether heterozygous SNPs (double peaks representing mixed infections) were identified using the heterozygous call plug-in tool in Geneious software package (Biomatters, Inc., San Francisco, CA) with a minor allele threshold of at least 30%. Mixed infections were recorded in each study except for Tanzania. For samples with mixed infections and SNP variations at codons 86, 184 and 1246, each possible haplotype constructed from the observed SNPs was reported for *Pfmdr1*(23)*. Pfmdr1* codons 1034 and 1042 were not included in the statistical analysis because the vast majority exclusively carried the wild-type SNP.

Mixed infections at the codons of interest included N and Y at codon 86 (N/Y), Y and F at codon 184 (Y/F), and D and Y at codon 1246 (D/Y). For the odds ratio analysis, mixed infections were reclassified to one SNP per codon for each treatment arm. This baseline SNP served as the exposure. For patients treated with AL, mixed infections at codon 86 were reclassified as N86; at codon 184 as 184F; and at codon 1246 as D1246. This is because previous studies have found N86, 184F, and D1246 to predominate in patients after AL treatment (9,11,12). For patients treated with ASAQ or DP, mixed infections at codon 86 were reclassified as 86Y; at codon 184 as Y184; and at codon 1246 as 1246Y. This is because previous studies have found 86Y, Y184, and 1246Y to predominate in patients after treatment with amodiaquine (11,13,24). Patients with mixed infections following treatment with DP were classified similarly to those treated with ASAQ because piperaquine has been shown to exert selective pressure on *Pfmdr1* in the same direction as amodiaquine (25,26). We examined all recurrent infections, which included recrudescent and new infections, rather than examining solely recrudescent infections. New infections were included in this analysis because the *Pfmdr1* SNPs present post-treatment could make the parasite less susceptible to ACTs since the parasite was able to re-infect individuals soon after treatment, negating the potential post-treatment prophylactic effect of the partner drug.

### Statistical analysis

All statistical analyses were conducted using R version 4.2.2 (R Foundation for Statistical Computing, Vienna, Austria). Multiple sensitivity analyses using only recrudescent infections were done to assess potential bias that could occur when analyzing new infections and recrudescent infections together. The first sensitivity analysis investigated the association between those treated with AL that experienced a recrudescent infection and *Pfmdr1* codons 86 and 184. This could not be done with *Pfmdr1* codon 1246 due to small sample size. A second sensitivity analysis investigated the association between those treated with ASAQ who experienced a recrudescent infection and *Pfmdr1* codon 184. Due to the limited sample size, a sensitivity analysis for those treated with ASAQ who experienced a recrudescent and *Pfmdr1* codon 86 and 1246 could not be conducted. Similarly, a sensitivity analysis was not performed for those treated with DP since so few recrudescent infections occurred in those receiving this ACT. After this, to determine whether patients infected with parasites carrying specific *Pfmdr1* SNPs were at increased risk of recurrent (recrudescent or new) infections, the Mantel-Haenszel (MH) method was used to create forest plots. To ensure there was not significant heterogeneity among studies, a Cochran’s Q test was used (*p* > 0.05). Since no statistically significant heterogeneity was found among studies, the common effect odds ratio was reported (27). Each forest plot was created to display the common effect odds ratio by treatment arm for each country-specific study and the pooled MH odds ratio by treatment arm when the data from all studies were compiled.

The common effect odds ratio represented the odds of a specific SNP in recurrent infections (in day of failure samples from those with recrudescent or new infections) compared to the odds of that same SNP in successfully treated infections (in pre-treatment samples from those who experienced an adequate clinical or parasitological response or in pre-treatment samples who initially cleared their infection but would later acquire a new infection). For the category of samples from successfully treated patients, we included: 1) pre-treatment samples from those with an adequate clinical and parasitological response; and 2) pre-treatment samples from those who would go on to be infected with a different parasite (i.e., new infection). This is because the pre-treatment infection was successfully cleared. For the category of unsuccessfully treated patients, we included: 1) the post-treatment recrudescent samples; and 2) the post-treatment samples from those patients who were newly infected (because the patient experienced an infection while the long-acting partner drug was still circulating). Conditional logistic regression was used to investigate the relationship between all the two-codon (e.g., 86Y and Y184) and three-codon haplotype (e.g., N86, 184F, and D1246) combinations with the occurrence of recurrent infections.

In patients who experienced either a recrudescent or new infection during the follow-up period, the *Pfmdr1* SNPs of the pre-treatment and recurrent sample were compared for each of these outcomes using McNemar’s test to examine selection. SNPs from pre-treatment samples were compared to their corresponding recurrent infection SNP to confirm that differences represented selection rather than original differences in SNP frequencies among the population (9).

## Results

### Baseline prevalence of genetic markers associated with resistance

The baseline prevalence of SNPs was determined using all successfully sequenced pre-treatment samples. Pre-treatment SNPs at position 86 were analyzed for 2,999 samples, at position 184 for 3,050 samples, at position 1034 for 3,006 samples, at position 1042 for 3,006 samples, and at position 1246 for 3,027 samples (Table 1).

**Table 1.**
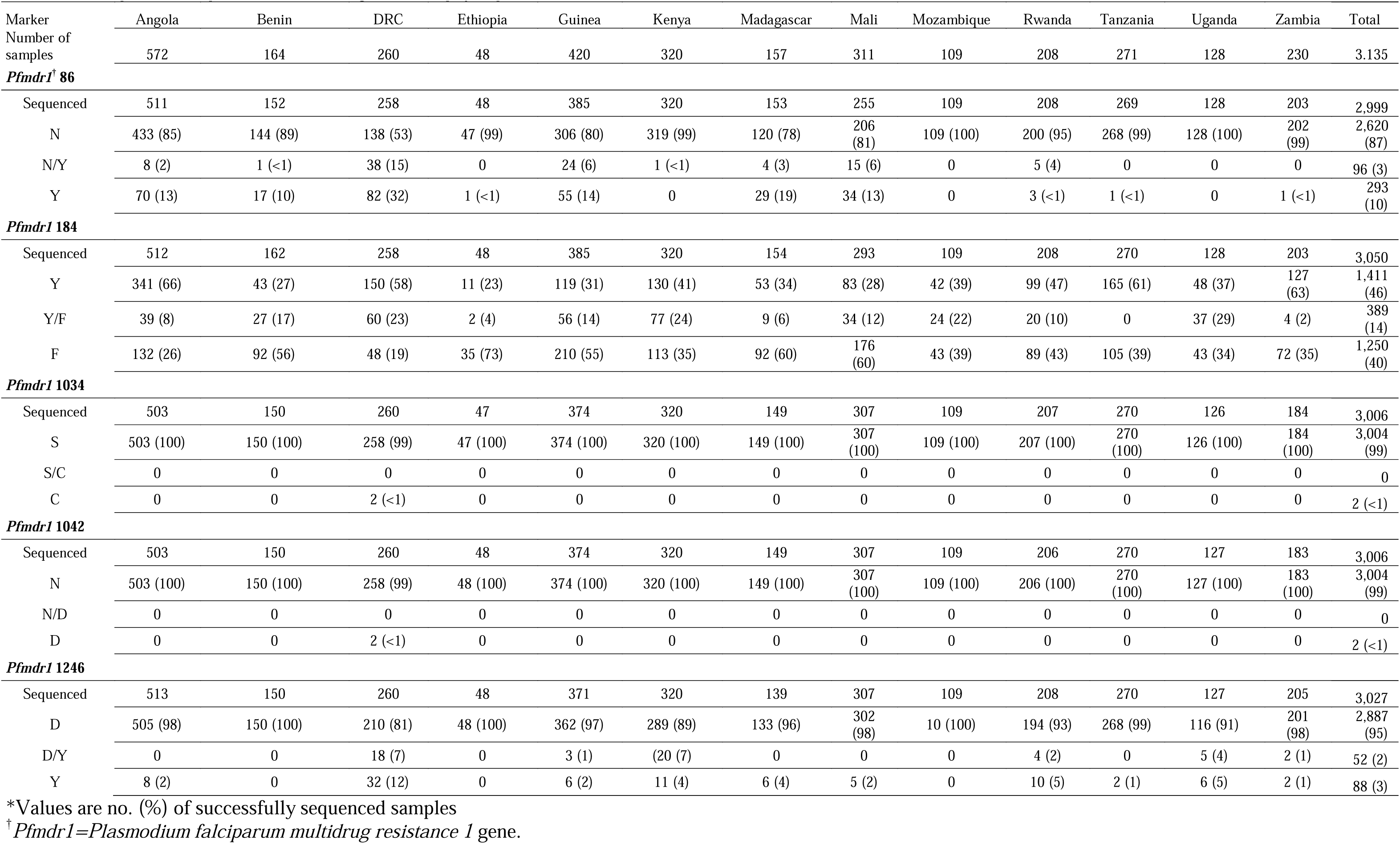
Baseline (pre-treatment) prevalence of *Pfmdr1* single nucleotide polymorphisms*.

The prevalence of *Pfmdr1* SNPs varied by country (Table 1). The *Pfmdr1* N86 SNP was found in the majority of samples from every country, with the prevalence ranging from 53– 100%. Samples from Ethiopia, Kenya, Mozambique, Tanzania, Uganda, and Zambia almost exclusively carried the *Pfmdr1* N86 SNP. *Pfmdr1* 184 had the most variation, with the prevalence of 184Y SNP ranging from 23–66%. Almost all samples carried parasites with the *Pfmdr1* S1034 and *Pfmdr1* N1042 wild-type alleles except for two participants from the Democratic Republic of the Congo, who carried parasites with both *Pfmdr1* 1034C and *Pfmdr1* 1042D. *Pfmdr1* D1246 SNP was found in 81–100% of the samples from most countries, with the Democratic Republic of the Congo having *Pfmdr1* D1246 in 81% of samples.

### Parasite single nucleotide polymorphisms as risk factors for recurrent infection

For patients treated with AL, the odds ratio between those with wild-type N86 in recurrent infections and those with wild-type N86 in infections that were successfully cleared was 5.22 (95% CI 2.69, 10.13) (Figure 2, panel a). For those treated with ASAQ, the odds of the 86Y SNP in recurrent infections was 2.72 (95% CI 1.28, 5.79) times higher than the odds of the same SNP in infections that were successfully cleared (Figure 2, panel b). There was no association detected between *Pfmdr1* codon 86 and recurrent infection for those treated with DP (Figure 2, panel c). Similarly, there was no association between *Pfmdr1* codons 184 or 1246 and recurrent infection in those treated with AL, ASAQ, or DP (Figures 3 and 4). When examining recrudescent infections alone (and not combined with new infections), the results of the sensitivity analysis were not statistically significant, although the odds ratios were either in the same direction as the aforementioned results or no effect was found (Supplementary Figures 1 and 2).

**Figure 2.**
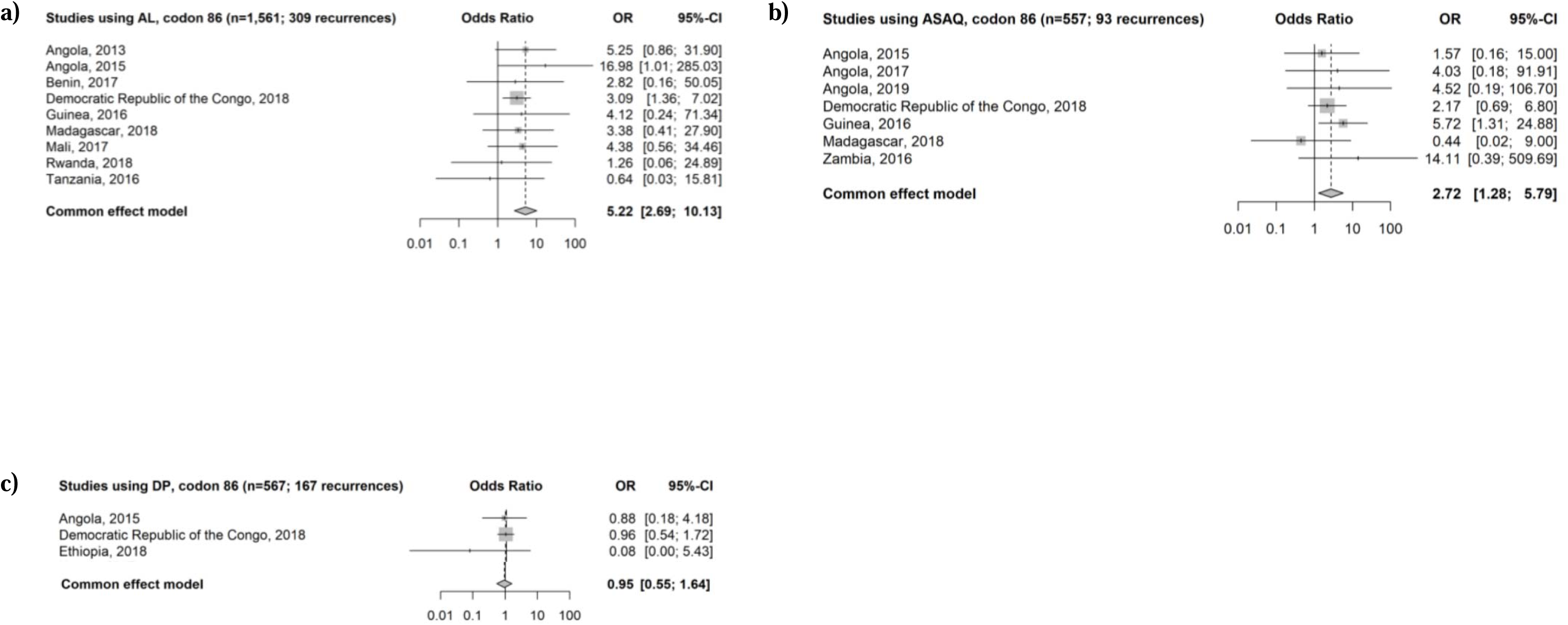
**a)** Forest plot of recurrent parasitemia in patients treated with AL. Odds ratio of N86 among patients with recurrent parasitemia compared to the odds ratio of N86 among patients who successfully cleared the infection. **b)** Forest plot of recurrent parasitemia in patients treated with ASAQ. Odds ratio of 86Y among patients with recurrent parasitemia compared to the odds ratio of 86Y among those who successfully cleared the infection. **c)** Forest plot of recurrent parasitemia in patients treated with DP. Odds ratio of 86Y among patients with recurrent parasitemia compared to the odds ratio of 86Y among patients who successfully cleared the infection. AL, artemether-lumefantrine; ASAQ, artesunate-amodiaquine; DP, dihydroartemisinin-piperaquine; OR, odds ratio; CI, confidence interval.

**Figure 3.**
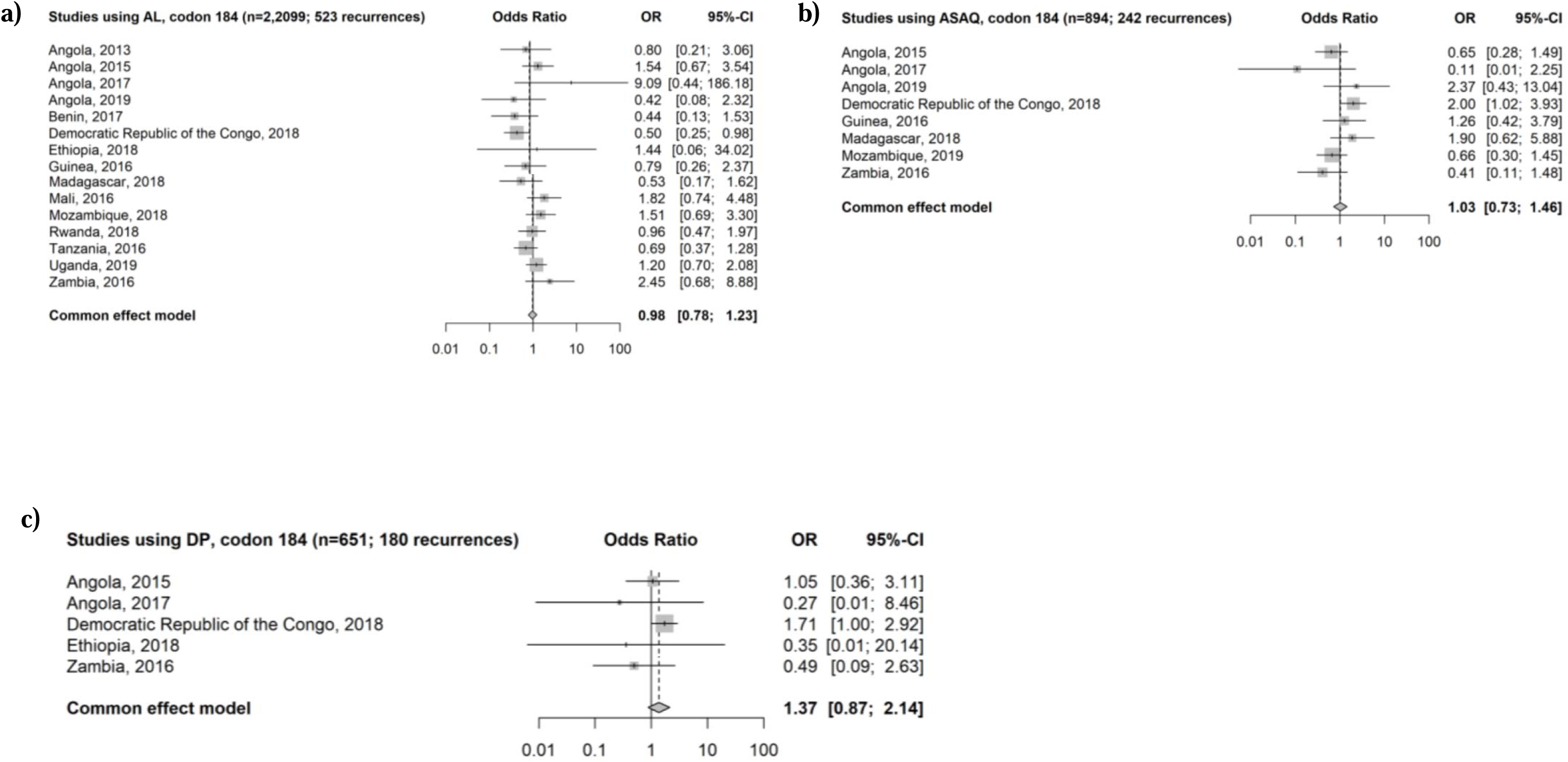
**a)** Forest plot of recurrent parasitemia in patients treated with AL. Odds ratio of 184F among patients with recurrent parasitemia compared to the odds ratio of 184F among patients who successfully cleared the infection. **b)** Forest plot of recurrent parasitemia in patients treated with ASAQ. Odds ratio of Y184 among patients with recurrent parasitemia compared to the odds ratio of Y184 among patients who successfully cleared the infection. **c)** Forest plot of recurrent parasitemia in patients treated with DP. Odds ratio of 86Y among patients with recurrent parasitemia compared to the odds ratio of Y184 among patients who successfully cleared the infection. AL, artemether-lumefantrine; ASAQ, artesunate-amodiaquine; DP, dihydroartemisinin-piperaquine; OR, odds ratio; CI, confidence interval.

**Figure 4.**
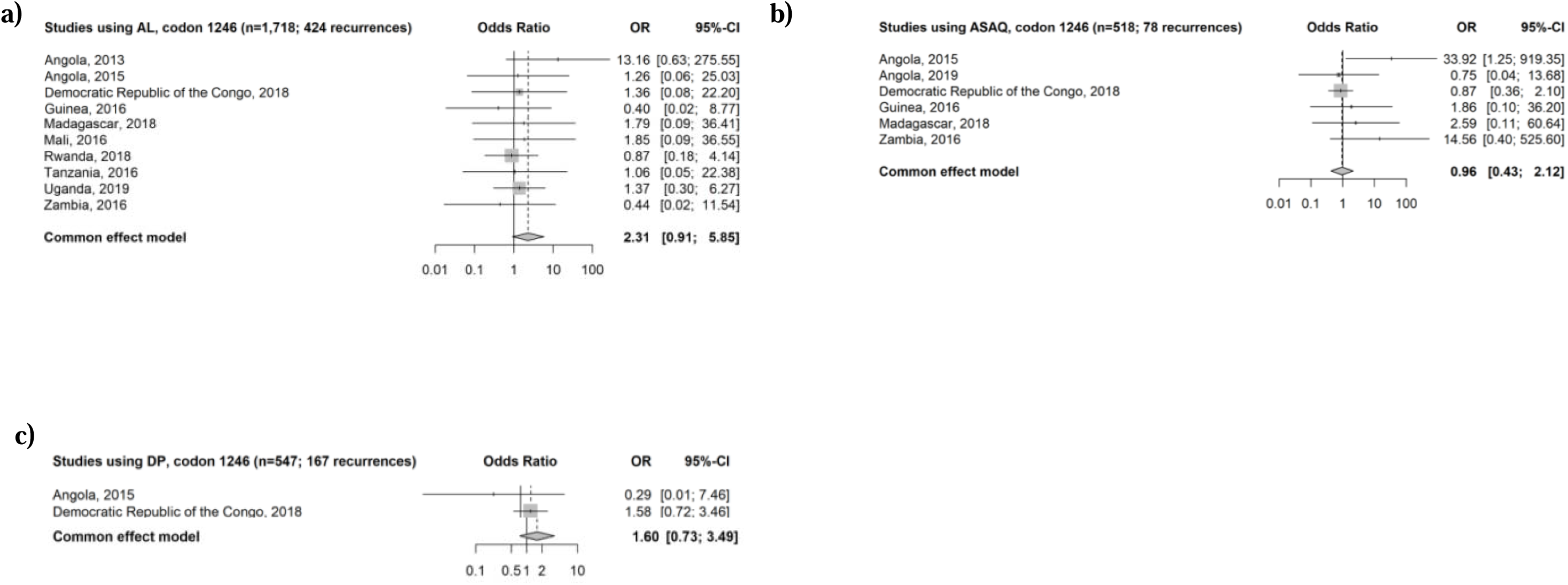
**a)** Forest plot of recurrent parasitemia in patients treated with AL. Odds ratio of D1246 among patients with recurrent parasitemia compared to the odds ratio of D1246 among patients who successfully cleared the infection. **b)** Forest plot of recurrent parasitemia in patients treated with ASAQ. Odds ratio of 1246Y among patient with recurrent parasitemia compared to the odds ratio of 1246Y among patients who successfully cleared the infection. **c)** Forest plot of recurrent parasitemia in patients treated with DP. Odds ratio of 1246Y among patients with recurrent parasitemia compared to the odds ratio of 1246Y among patients who successfully cleared the infection. AL, artemether-lumefantrine; ASAQ, artesunate-amodiaquine; DP, dihydroartemisinin-piperaquine; OR, odds ratio; CI, confidence interval.

Country-specific odds ratios could not be generated for several countries for some treatment arms and SNPs. For example, patients who received AL in the Uganda TES only had parasites carrying the N86 SNP and none carrying the 86Y SNP following reclassification of mixed infections. Thus, the country-specific odds ratio was undefined and could not contribute to the overall, pooled MH odds ratio.

### Post-treatment selection of genetic markers associated with reduced susceptibility

To examine changes in the genetic markers of parasites after drug treatment during the period of follow-up, we compared the prevalence of *Pfmdr1* SNPs in paired isolates from the initial and the recurrent parasites (Table 2). Statistically significant selection of *Pfmdr1* N86 occurred in new infections as well as recurrent infections (recrudescent infections plus new infections) after treatment with AL. There was no statistically significant selection of *Pfmdr1* SNPs at codons 184 or 1246 after AL treatment and no statistically significant selection of SNPs at *Pfmdr1* codons 86, 184, or 1246 after ASAQ or DP treatment.

**Table 2.** Selection of *Pfmdr1* genotypes after treatment with artemether-lumefantrine, artesunate-amodiaquine, and dihydroartemisinin-piperaquine*.

| Marker | Genotype<sup>†</sup> | Recurrent infection |  |  | □ | Recrudescent infection |  |  | □ | New infection |  |  |
| --- | --- | --- | --- | --- | --- | --- | --- | --- | --- | --- | --- | --- |
|  |  | AL | ASAQ | DP |  | AL | ASAQ | DP |  | AL | ASAQ | DP |
| *Pfmdr1* 86 | N→Y | (11/647) | (9/99) | (22/205) | □ | (1/153) | (1/27) | (2/37) | □ | (10/494) | (8/72) | (20/168) |
| □ | Y→N | (32/647) | (22/99) | (26/205) | □ | (3/153) | (5/27) | (3/37) | □ | (29/494) | (17/72) | (23/168) |
| □ | No change** | (604/647) | (68/99) | (157/205) | □ | (149/153) | (21/27) | (32/37) | □ | (455/494) | (47/72) | (125/168) |
| P value | □ | **<0.01** | 0.03<br>(exact) | 0.56 | □ | 0.62 | 0.22<br>(exact) | 1<br>(exact) | □ | **<0.01** | 0.11 (exact) | 0.65 |
| *Pfmdr1* 184 | Y→F | (119/639) | (17/101) | (44/205) | □ | (22/153) | (15/36) | (7/37) | □ | (95/486) | (10/74) | (39/168) |
| □ | F→Y | (117/639) | (16/101) | (35/205) | □ | (27/153) | (2/36) | (5/37) | □ | (92/486) | (15/74) | (28/168) |
| □ | No change** | (403/639) | (68/101) | (126/205) | □ | (104/153) | (19/36) | (25/37) | □ | (299/486) | (49/74) | (101/168) |
| P value | □ | 0.89 | 1 | 0.31 | □ | 0.48 | 0.29<br>(exact) | 0.77<br>(exact) | □ | 0.82 | 0.42 (exact) | 0.18 |
| *Pfmdr1* 1246 | D→Y | (9/618) | (6/105) | (16/205) | □ | (2/150) | (2/27) | (2/37) | □ | (7/468) | (6/78) | (14/168) |
| □ | Y→D | (17/618) | (8/105) | (9/205) | □ | (2/150) | (2/27) | (2/37) | □ | (15/468) | (4/78) | (7/168) |
| □ | No change** | (592/618) | (91/105) | (180/205) | □ | (146/150) | (23/27) | (33/37) | □ | (446/468) | (68/78) | (147/168) |
| P value | □ | 0.17 | 0.77<br>(exact) | 0.16 | □ | 1 | 1 (exact) | 1<br>(exact) | □ | 0.14 | 0.75 (exact) | 0.19 |

AL, artemether-lumefantrine; ASAQ, artesunate-amodiaquine; DP, dihydroartemisinin-piperaquine
\*Values in bold indicate statistically significant selection (P<0.05) using McNemar's paired test. Those marked exact were tested by using the exact distribution for sample sizes less than 100.
*Pfmdr1*=*Plasmodium falciparum* multidrug resistance 1 gene<sup>†</sup>Each category includes all changes from one allele to another. For example, N→Y includes N→Y, N→N/Y, and N/Y→Y changes; Y→N includes Y→N, Y→N/Y, N/Y→N changes.
\*\*No change in genotype from before and after treatment.
Note: recurrent infections are the combination of recrudescent and new infections

### Parasite haplotypes as a risk factor for recurrent infection

Recurrent infections, which included recrudescent and new infections, were pooled to assess the relationship between *Pfmdr1* haplotype and recurrent infections. Only one haplotype, N86/184F, was found to be associated with recurrent infection. The odds of N86/184F among individuals with recurrent infection after AL treatment (post-treatment) was 6.36 (95% CI 1.39, 29.19) times higher than the odds of the same haplotype among those that successfully cleared the infection (pre-treatment). The combined effect of carrying N86/184F had a greater measure of association (OR: 6.36, 95% CI: 1.39, 29.19) than recurrent parasites only carrying N86 (OR: 5.22, 95% CI: 2.69, 10.13). There was no statistically significant selection of *Pfmdr1* haplotypes at codons 184 or 1246 after AL treatment and no statistically significant selection of haplotypes at *Pfmdr1* codons 86, 184, or 1246 after ASAQ or DP treatment.

## Discussion

This pooled analysis of data from 16 efficacy studies suggests that SNPs in *Pfmdr1* codon 86 influence recurrent infection after AL and ASAQ treatment. Individuals infected with parasites that carried the *Pfmdr1* N86 SNP were statistically more likely to experience a recurrent infection after treatment with AL. This finding is consistent with a previous study which found that patients treated with AL and infected with parasites carrying the *Pfmdr1* N86 allele were at statistically greater risk of treatment failure than those whose parasites carried 86Y(9). This increased risk of recurrent infection, in addition to significant selection of *Pfmdr1* N86 by AL, could be an indicator of decreased susceptibility to the partner drug, lumefantrine.

For our haplotype analysis, patients given AL and infected with parasites carrying *Pfmdr1* N86/184F were statistically more likely to cause a new infection. This finding is similar to a study which found that after AL treatment, individuals carrying *Pfmdr1* N86/184F/D1246 were able to re-infect with lumefantrine blood concentrations 15-fold higher than those with the 86Y/Y184/1246Y haplotype (8). Another study discovered that individuals with *Pfmdr1* N86/D1246 experienced a new infection earlier after AL treatment (9). These findings suggest that monitoring for decreasing post-treatment durations until a new infection has occurred or surveilling for the prevalence of certain post-treatment *Pfmdr1* haplotypes may also provide indications of decreased susceptibility to lumefantrine.

After treatment with ASAQ, individuals infected with parasites that carried the *Pfmdr1* 86Y SNP were statistically more likely to cause a recurrent infection, shown in this study. Like our results from those treated with AL, a high prevalence of *Pfmdr1* 86Y SNP may be an early warning signal of resistance to the partner drug, amodiaquine, but this association requires further investigation because prior studies are collectively inconclusive. An *in vitro* analysis from Southern Sudan found a significant association between failure after amodiaquine treatment and the combined presence of *Pfmdr1* 86Y and *Pfcrt* 76T but no association with *Pfmdr1* 86Y alone (28). However, an *in vivo* study from Burkina Faso found a significant association between *Pfmdr1* 86Y and treatment failures after amodiaquine treatment and another *in vivo* study from Burkina Faso showed significant selection of *Pfmdr1* 86Y after treatment with ASAQ (24,29).

Additionally, a meta-analysis revealed that the risk of therapeutic failure was greater for participants who carried *Pfmdr1* 86Y and were treated with amodiaquine monotherapy (30). Our results align with the findings from Burkina Faso and the meta-analysis. Close monitoring of the prevalence of *Pfmdr1* codon 86 before and after treatment with amodiaquine monotherapy or ASAQ could be used to indicate potential partner-drug resistance to amodiaquine.

Unlike previous studies, this analysis did not find statistically significant selection of *Pfmdr1* SNPs by ASAQ (9,11,31–34). There were no significant findings between the use of DP and treatment outcomes or selection of *Pfmdr1* SNPs by DP. While this lack of association has been found before, there has also been evidence of piperaquine exerting a selective pressure on *Pfmdr1* genes similar to amodiaquine (25,26). However, there was clear evidence of significant selection of *Pfmdr1* N86 by AL in those who experienced either a recurrent infection (i.e., recrudescent or new infection) or a new infection alone in this study. This result aligns with prior studies that found selection of particular *Pfmdr1* SNPs among parasite populations after treatment with AL (8,9,12,15,17,35–40). Additionally, the significant selection of *Pfmdr1* N86 by AL is consistent with prior molecular studies that identified changes in the parasite population prevalence of *Pfmdr1* SNPs after the introduction or greater use of lumefantrine (14,15,34–37,41,42).

Although SNPs at *Pfmdr1* codon 86 were associated with recurrent infections (i.e., recrudescent or new infections), they were not associated solely with recrudescent infections in those treated with AL or ASAQ, an association shown in a prior study (9). The small number of recrudescent infections, a limitation of this study even with pooled data across 16 TES, may have prevented this analysis from finding significant results. A similar limitation was the smaller number of patients receiving ASAQ, which may have contributed to the lack of evidence of selection for *Pfmdr1* 86Y by ASAQ, a selection other studies have demonstrated [16, 23, 49–51]. Another limitation was that information on the number of *Pfmdr1* copies was not available for the majority of studies included in this analysis but has been previously found as a predictor of treatment failure after AL treatment (10,17). Additionally, most of the efficacy studies enrolled patients under 6 years old. Therefore, our data may not be applicable to adult populations. Lastly, efficacy studies are not countrywide but occur in just a few sites, which may not be representative of an entire country’s parasite population. Despite its limitations, our study possessed many strengths, including the large number of samples from numerous countries that were sequenced at the same laboratory using the same methodology.

Two patients from the Democratic Republic of the Congo carried parasites bearing the *Pfmdr1* haplotype commonly found in South America, *Pfmdr1* 1034C, 1042D, and 1246Y, which has been associated with increased sensitivity to artemisinin (39,43,44). However, in our study, the participants experienced a new infection after treatment with DP. Because detailed travel histories were not available for these patients, it is unknown whether they acquired their infections locally or abroad.

Changes in the prevalence of *Pfmdr1* SNPs over multiple TESs can be a subtle sign of selection of parasite populations by AL or ASAQ. Decreasing efficacy of lumefantrine, amodiaquine, or other partner drugs leaves the artemisinin component of an ACT vulnerable to new selective pressures. Modeling demonstrates that partner drug resistant *Pfmdr1* genotypes may lead to establishment of artemisinin resistance (45). This phenomenon may already be occurring in Africa. Rwanda and Uganda, two countries reporting *K13* mutations associated with artemisinin resistance, use AL as their first-line ACT and had a predominance of the *Pfmdr1* N86 genotype in our study (7,46,47). Previous studies have shown that ASAQ and AL exert opposing selecting pressures on *Pfmdr1* 86 (9). Depending on the prevalence of *Pfmdr1* polymorphisms, the opposing selection by the partner drugs could better inform which ACT national malaria control programs could choose as a first-line therapy to delay resistance to both drugs. New strategies, such as multiple first-line therapies or dosing lumefantrine and amodiaquine together as part of a triple therapy, may overcome the negative effects of a particular *Pfmdr1* SNPs associated with decreased susceptibility to partner drugs, although that remains to be proven (48–50). Monitoring prevalence of *Pfmdr1* codons, such as N86Y, may indicate a parasite genetic background conferring reduced susceptibility to a particular antimalarial.

## Data Availability

All data produced in the present study are available upon reasonable request to the authors.

## Acknowledgements

The authors would like to acknowledge Venkatachalam Udhayakumar for his suggestions on the manuscript and for supporting the laboratory genotyping trainings vital to this multi-country endeavor. All authors have no conflicts of interest.

## Disclaimer

The opinions expressed herein are those of the author(s) and do not necessarily reflect the views of any collaborating institute including the Centers for Disease Control and Prevention or the US Agency for International Development.

## Author contributions

Developed the study idea: VL, MP, MV, EH

Collected the data: MR, MA, HW, DI, AC, PD, AA, AB, SK, SN, AU, GK, AA, OK, NL, SS, ZZ

Analyzed the data: VL, MP, KR, LM, EH

Wrote the manuscript: VL, MP, EH

Approved the manuscript: All

## Supplementary

**Supplementary Table 1.**
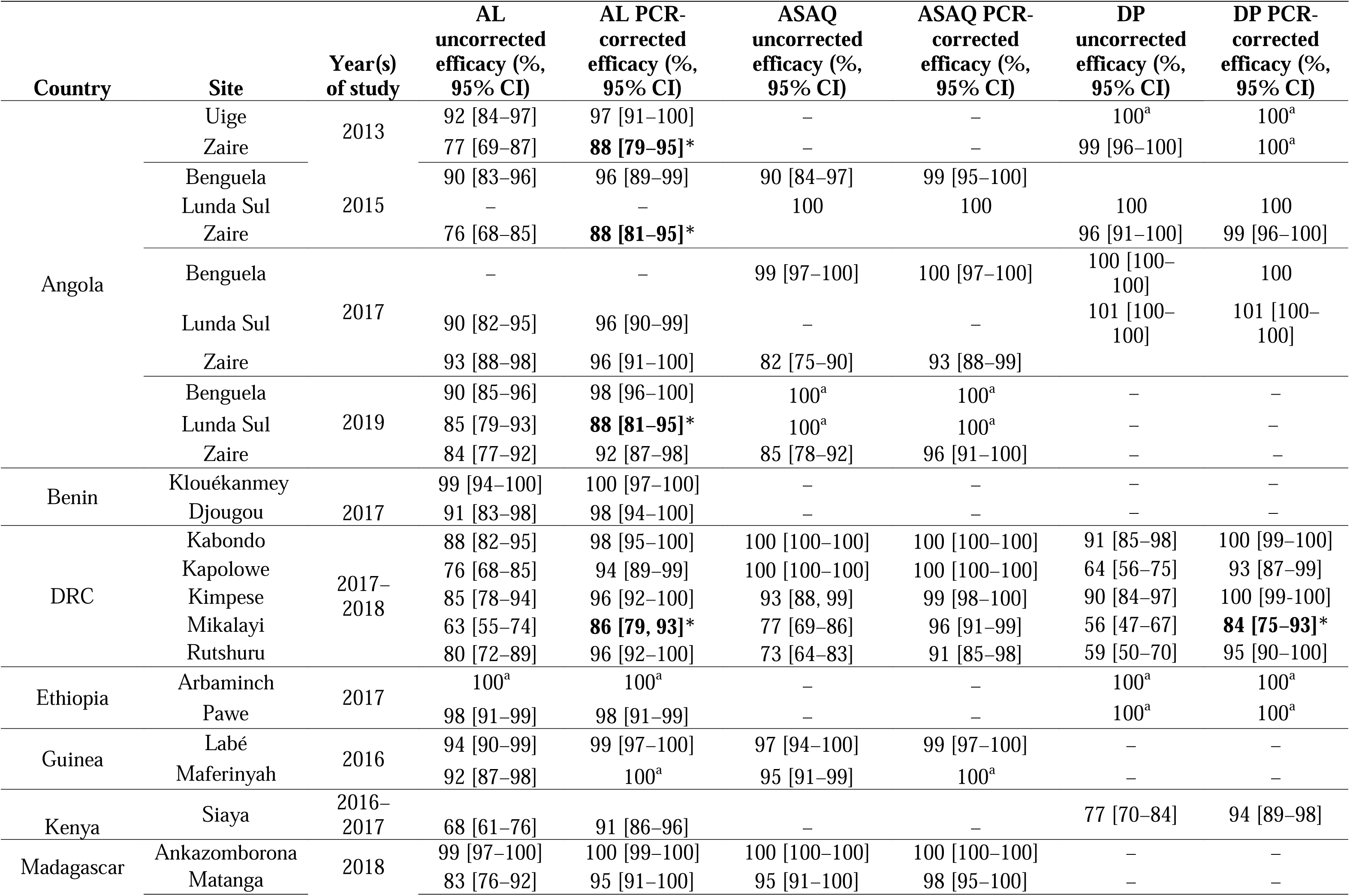
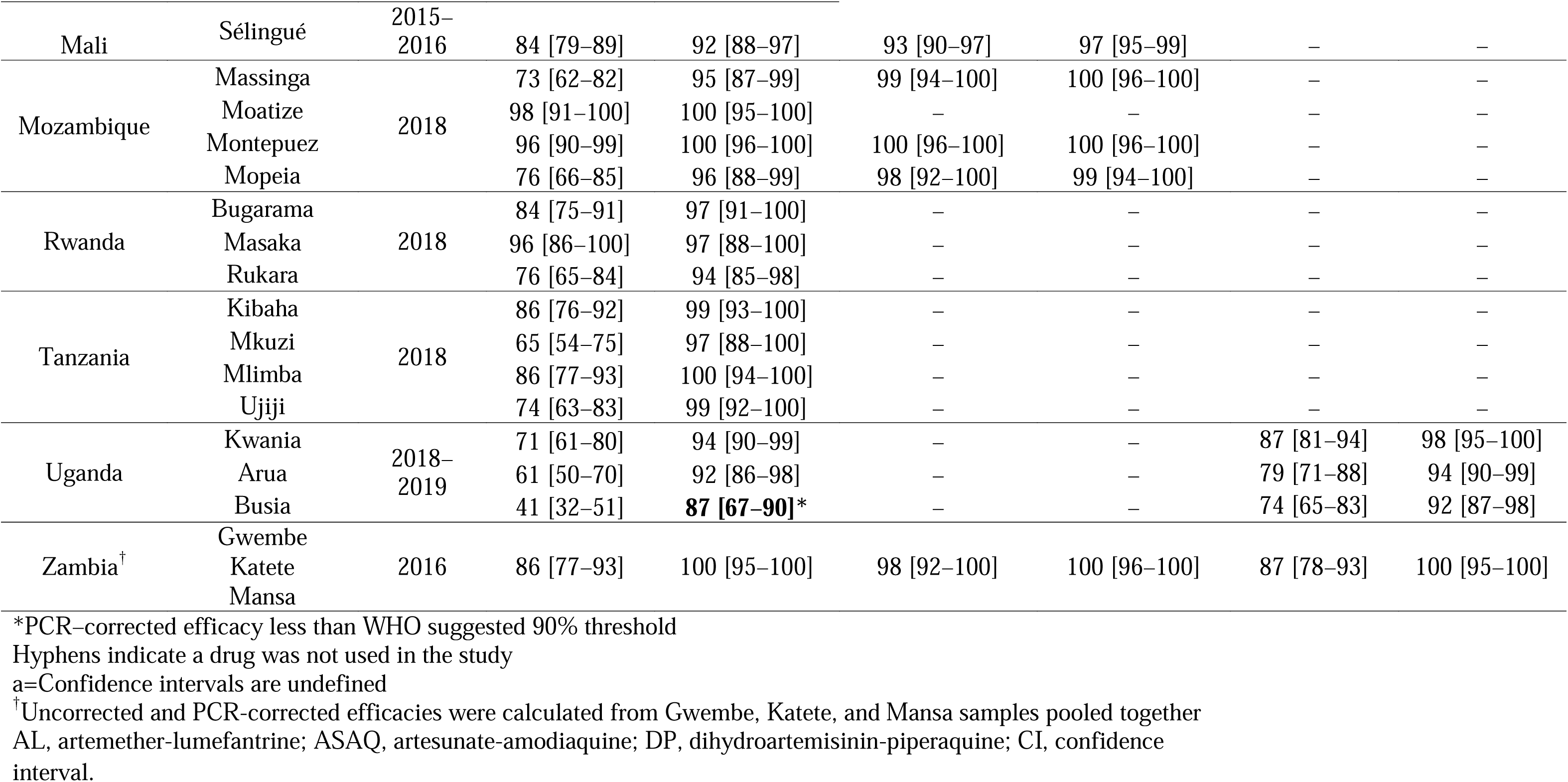
Uncorrected and PCR–corrected efficacies for therapeutic efficacy study sites.

**Supplementary Table 2.** Summary of antimalarial therapeutic efficacy studies, thirteen countries in Africa, 2013–2019.

| Country | Sites | ACT(s) studied* | Study year(s) | Patient age | Number of samples | Number of patients who completed study | Genotyping method | Publication |
| --- | --- | --- | --- | --- | --- | --- | --- | --- |
| Angola | Uige, Zaire | AL, DP | 2013 | 6 mo–9 y | 50 | 320 | Microsatellites | (1) |
|  | Benguela, Lunda Sul, Zaire | AL, ASAQ, DP | 2015 | 6 mo–12 y | 467 | 467 | Microsatellites | (2,3) |
|  | Benguela, Lunda Sul, Zaire | AL, ASAQ, DP | 2017 | 6 mo–12 y | 76 | 540 | Microsatellites | (4) |
|  | Benguela, Lunda Sul, Zaire | AL, ASAQ | 2019 | 6 mo–12 y | 106 | 579 | Microsatellites | (5) |
| Benin | Klouékanmey, Djougou | AL | 2017 | 6 mo–59 mo | 175 | 235 | Microsatellites | Country report |
| DRC | Kabondo, Kapolowe, Kimpese, Mikalayi, Rutshuru | AL, ASAQ, DP | 2017–2018 | 6 mo–59 mo | 520 | 1,271 | Microsatellites | (6) |
| Ethiopia | Arba Minch, Pawe | AL, DP | 2017 | >6 mo | 50 | 170 | Microsatellites | (7) |
| Guinea | Labé, Maferinyah | AL, ASAQ | 2016 | 6 mo–59 mo | 443 | 413 | Microsatellites | (8) |
| Kenya | Siaya | AL, DP | 2016–2017 | 6 mo–59 mo | 415 | 270 | *msp1*, *msp2*, *glurp* | (9, 10) |
| Madagascar | Ankazomborona, Antensenavolo, Kianjavato, Matanga, Vohitromby | AL, ASAQ | 2018 | 6 mo–14 y | 179 | 335 | Microsatellites | (11) |
| Mali | Sélingué | AL, ASAQ | 2015–2016 | 6 mo–59 mo | 386 | 434 | *msp1*, *msp2*, *glurp* | (12) |
| Mozambique | Massinga, Moatize, Montepuez Mopeia | AL, ASAQ | 2018 | 6 mo–59 mo | 158 | 641 | Microsatellites | (13, 14) |
| Rwanda | Bugarama, Masaka, Rukara | AL | 2018 | 6 mo–59 mo | 243 | 224 | Microsatellites | (15) |
| Tanzania | Kibaha, Mkuzi, Mlimba, Ujiji | AL | 2016 | 6 mo–10 y | 339 | 335 | Microsatellites | (16) |
| Uganda | Aduku, Arua, Masafu | AL, DP | 2018–2019 | 6 mo–10 y | 256 | 590 | Microsatellites | (17) |
| Zambia | Gwembe, Katete, Mansa | AL, ASAQ, DP | 2016 | >6 mo | 266 | 291 | Microsatellites | Country report |

\*ACTs, artemisinin-based combination therapies; AL, artemether-lumefantrine; ASAQ, artesunate-amodiaquine; DP, dihydroartemisinin-piperaquine; *msp1*, merozoite surface protein-1; *msp2*, merozoite surface protein-2; *glurp*, glutamate-rich protein; DRC, Democratic Republic of the Congo

**Supplementary Figure 1.**
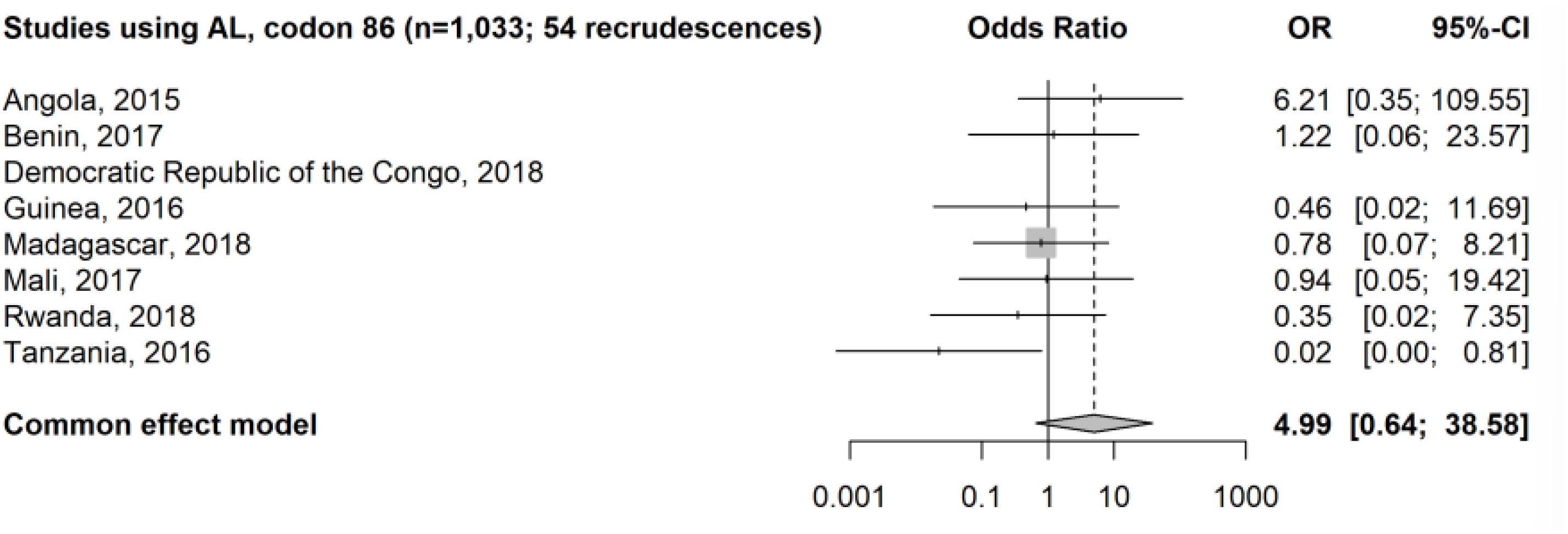
Forest plot of recrudescent infection in patients treated with AL. Odds ratio of N86 among patients with recrudescent infection compared to the odds ratio of N86 among patients who successfully cleared the infection. OR, odds ratio; CI, confidence interval.

**Supplementary Figure 2.**
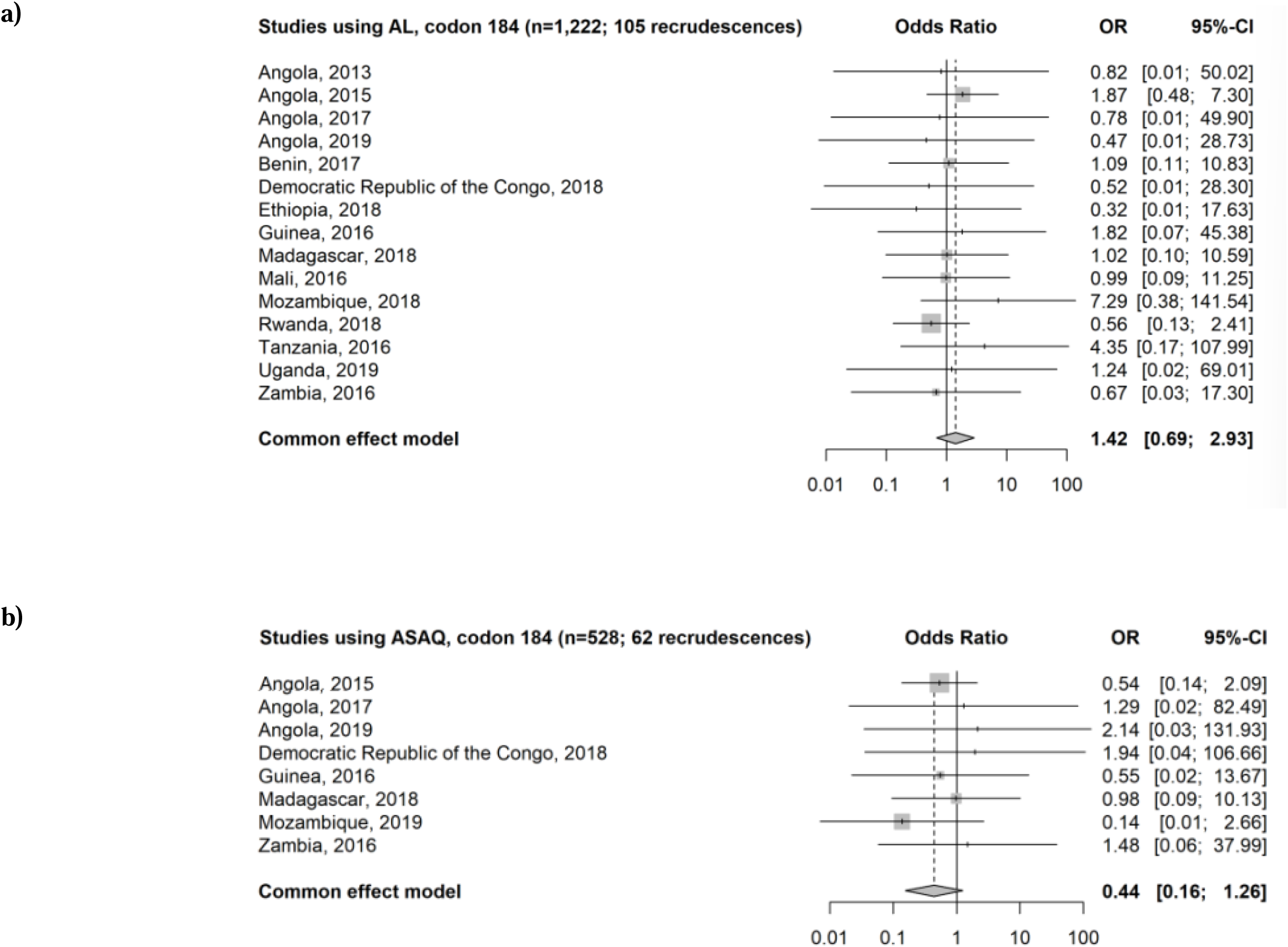
**a)** Forest plot of recrudescent infection in patients treated with AL. Odds ratio of 184F among patients with recrudescent infection compared to the odds ratio of 184F among patients who successfully cleared the infection. **b)** Forest plot of recrudescent infection in patients treated with ASAQ. Odds ratio of Y184 among patients with recrudescent infection compared to the odds ratio of Y184 among patients who successfully cleared the infection. OR, odds ratio; CI, confidence interval.

